# Falsification Testing of Sepsis Prediction Models: Evaluating Independent Biological Signal After Controlling for Care-Process Intensity

**DOI:** 10.64898/2026.03.17.26348414

**Authors:** Adam Dickens

## Abstract

**Background:** Automated sepsis early-warning systems have attracted substantial research investment, yet a fundamental question remains unresolved: do these models detect independent biological signals, or do they predominantly learn care-process intensity — the pattern of clinician ordering behavior applied to patients already suspected of being ill? We report a pre-registered falsification study testing this hypothesis across four independent clinical datasets.

**Methods:** A four-phase falsification framework with pre-specified thresholds was registered on OSF (March 11, 2026) before any data access. The primary confirmatory analysis used MIMIC-IV v3.1 (n=65,241 adult ICU stays, Beth Israel Deaconess Medical Center, 2008-2022). Exploratory replication analyses used eICU-CRD v2.0 (n=136,864, 208 US hospitals), MIMIC-III v1.4 (n=44,091), and the PhysioNet/CinC 2019 Sepsis Challenge (n=40,314). Each phase tested a distinct falsification criterion: (1) concordance across Sepsis-2, Sepsis-3, and CMS SEP-1 definitions; (2) model performance degradation when care-intensity proxy features are removed; (3) predictive performance of care-intensity features alone; and (4) discriminability of synthetic records generated to match care-intensity distributions.

**Results:** The pre-registered primary analysis (MIMIC-IV) did not confirm the hypothesis (0/4 phases confirmed). Biological features predicted Sepsis-3 labels with AUROC 0.901 (95% CI 0.899-0.904); removing care-intensity features reduced performance by only 0.003 AUROC (drop=0.0027). The pre-specified Phase 3 threshold (care-only AUROC >0.70) was not met by the primary logistic regression model (AUROC 0.660); however, a sensitivity XGBoost model did exceed the threshold (AUROC 0.729), suggesting nonlinear care-intensity signal. However, a clinically significant finding emerged consistently across all four datasets: mean pairwise Jaccard similarity between clinical sepsis definitions and administrative coding (CMS SEP-1) was approximately 0.32 at the primary site and 0.20 across multi-center cohorts, indicating that hospital quality metrics and regulatory reporting systematically measure a different patient population than clinical definitions identify. Exploratory analyses revealed a detectable care-intensity signal in the eICU multi-center cohort (AUC drop=0.076) not present at the single academic center. At community hospitals the care-intensity signal was larger (AUC drop 0.076 versus 0.003 at the academic center), suggesting the null result may be specific to high-acuity academic settings.

**Conclusions:** At an elite academic medical center, sepsis prediction models detect genuine biological signal. Care-process leakage is not the primary driver of model performance in MIMIC-IV. The more consequential and robust finding is the systematic divergence between clinical and administrative sepsis definitions across all datasets examined, which has direct implications for regulatory reporting, pay-for-performance metrics, and the validity of AI benchmarks built on administrative data.

## Introduction

Sepsis remains among the leading causes of ICU mortality, and automated early-warning systems based on electronic health record (EHR) data have attracted enormous research investment over the past decade. Models trained on large clinical databases such as MIMIC and eICU report AUROC values routinely exceeding 0.85, suggesting strong predictive performance. However, a fundamental methodological concern has not been rigorously evaluated: whether these models detect independent biological signals of sepsis, or whether they predominantly learn to recognize the pattern of intensive clinician ordering behavior applied to patients already suspected of being ill.

This distinction — between detecting biology and detecting clinician suspicion — has profound clinical implications. A model that identifies care-process intensity cannot provide early warning ahead of existing clinical suspicion; it can only confirm suspicion that already exists. Furthermore, models that learn care-intensity patterns risk amplifying existing clinician biases, flagging patients who receive intensive monitoring rather than those who are biologically deteriorating.

A second, distinct concern is the validity of sepsis outcome labels themselves. Three major definitional frameworks — Sepsis-2 (SIRS-based), Sepsis-3 (SOFA-based), and CMS SEP-1 (ICD code-based) — are used interchangeably in the literature despite representing conceptually distinct constructs. Administrative ICD coding serves as the basis for hospital quality reporting, regulatory metrics, and widely-cited US sepsis incidence estimates. Rhee et al. (2017) estimated approximately 1.7 million adult sepsis hospitalizations annually in the United States using clinical EHR criteria, noting that administrative code-based estimates diverge substantially from clinical ones. If these definitions identify substantially different patient populations, then models benchmarked against administrative codes may be measuring billing behavior rather than biological disease.

We report a pre-registered falsification study designed to test both concerns empirically. The study was registered on OSF (March 11, 2026) before any patient data was accessed. Pre-specified thresholds were established for each of four falsification phases. The primary confirmatory analysis was performed on MIMIC-IV v3.1. Three independent datasets served as exploratory replication cohorts. Results are reported transparently regardless of direction, consistent with the pre-registration commitment.

## Methods

### Study Design and Pre-Registration

This study employs a pre-specified falsification framework. The hypothesis, statistical thresholds, primary dataset, inclusion criteria, and analysis plan were registered on the Open Science Framework (OSF) at https://osf.io/9tbjm on March 11, 2026, prior to any access to patient data (CITI certification obtained; PhysioNet credentialing approved March 6, 2026, with Dr. Matthieu Komorowski serving as reference). The registration was timestamped and embargoed to prevent post-hoc modification. All deviations from the pre-registration are reported transparently in this manuscript.

### Datasets

The primary confirmatory analysis used MIMIC-IV Clinical Database v3.1 (PhysioNet), containing all adult ICU admissions at Beth Israel Deaconess Medical Center, Boston, MA from 2008-2022. Three exploratory replication datasets were analyzed after the primary results were obtained: eICU Collaborative Research Database v2.0 (208 US hospitals, variable years), MIMIC-III Clinical Database v1.4 (BIDMC, 2001-2012), and the PhysioNet/CinC 2019 Sepsis Challenge dataset (two hospital systems). All datasets were accessed under PhysioNet Data Use Agreements.

### Cohort Inclusion and Exclusion

Inclusion criteria applied uniformly across all datasets: adult patients (age >=18 years), first ICU stay per patient only (subsequent readmissions excluded), ICU length of stay >=4 hours, minimum one documented vital sign, and minimum one laboratory result. For eICU, the primary analysis unit was the unit stay; antibiotic and blood culture matching was performed at the hospital stay level (patienthealthsystemstayid) to account for multi-unit transfers, then mapped back to unit stay identifiers.

**Table 1.** Cohort summary across all four datasets.

| Dataset | N (Final Cohort) | Source | Period |
| --- | --- | --- | --- |
| MIMIC-IV v3.1 (Primary) | 65,241 | BIDMC, Boston MA | 2008-2022 |
| eICU-CRD v2.0 (Exploratory) | 136,864 | 208 US hospitals | Variable |
| MIMIC-III v1.4 (Exploratory) | 44,091 | BIDMC, Boston MA | 2001-2012 |
| PhysioNet/CinC 2019 (Exploratory) | 40,314 | 2 hospital systems | Variable |

### Sepsis Label Definitions

Three sepsis definitions were applied to each cohort where data permitted. Sepsis-2 required SIRS criteria >=2 (temperature <36 or >38 degrees C, heart rate >90/min, respiratory rate >20/min, WBC <4 or >12 x10^9/L) plus an infection proxy. Sepsis-3 required SOFA score >=2 plus the same infection proxy. CMS SEP-1 used ICD-9 codes 995.91, 995.92, 785.52 and ICD-10 codes beginning A40, A41, R65.2, or R57.2 (septic shock).

The infection proxy was defined permissively following the Sepsis-3 consensus approach: antibiotic prescription AND/OR blood culture order within a timing window (antibiotic <=72 hours after culture initiation, OR culture <=24 hours after antibiotic start). Antibiotics were identified by keyword match across 43 drug names in prescription records. Blood cultures were identified from microbiology events where specimen type contained ‘BLOOD’. The PhysioNet/CinC 2019 dataset does not include ICD codes; CMS SEP-1 was therefore not computable and Jaccard comparisons are reported only between Sepsis-2 and Sepsis-3.

SOFA scores were computed manually from chartevents, labevents, and inputevents where derived tables were unavailable. Components computed: respiratory (PaO2/FiO2 ratio, minimum per stay), coagulation (platelet count, mean per stay as a proxy for minimum — a conservative deviation from the Sepsis-3 specification which calls for the minimum value; this underestimates SOFA coagulation scores), liver (bilirubin, maximum per stay), cardiovascular (MAP minimum <70 mmHg plus vasopressor dose scoring from inputevents using published mcg/kg/min thresholds for norepinephrine, epinephrine, dopamine, dobutamine, vasopressin, and phenylephrine), CNS (minimum GCS total across stay, derived by pairing eye/verbal/motor components charted within a 5-minute window using a nearest-neighbor merge, then summing to obtain total GCS at each timepoint and taking the minimum across the stay; this approach corrects for components charted at slightly different timestamps), and renal (creatinine maximum). Maximum total SOFA per stay was used.

### Feature Construction

The full feature matrix included biological features and care-intensity proxy features. Biological features comprised: laboratory values (creatinine, glucose, lactate, WBC, hemoglobin, platelets, bilirubin, sodium, potassium — mean and maximum per stay), vital sign values (heart rate, systolic BP, MAP, respiratory rate, SpO2, temperature — mean per stay; SBP minimum), and demographics (age, gender). Additional biological features used in the production analysis (BUN, weight, and 30 Elixhauser comorbidity flags derived from ICD-10 codes) are not included in the public reproduction code for IP reasons; their inclusion does not materially affect the primary Phase 1 or Phase 2 findings, which are verified against SHA-256 checksums in the OSF proof package.

Care-intensity proxy features were four composite rates per ICU hour: lab ordering frequency (lab orders / LOS hours), vital measurement rate (vital sign measurements / LOS hours), nursing note frequency (datetimeevents count / LOS hours), and physician order rate (sum of inputevents, procedureevents, and prescriptions / LOS hours). Missing feature values were imputed with column medians computed across the analytic cohort prior to model training.

### Statistical Analysis

Phase 1 (Ground Truth Stability): Pairwise Jaccard similarity coefficients were computed for all three definitional pairs. Mean Jaccard was the primary metric. Pre-specified threshold: mean Jaccard < 0.50.

Phase 2 (Feature Dependence Test): XGBoost classifiers (n_estimators=300, max_depth=6, learning_rate=0.05, subsample=0.8, colsample_bytree=0.8) were trained on Sepsis-3 labels using 5-fold stratified cross-validation. Full feature model AUROC was compared to biological-feature-only model AUROC. Bootstrap 95% confidence intervals used 1,000 iterations with replacement. Pre-specified threshold: AUC drop > 0.15.

Phase 3 (Care-Intensity Universality): Logistic regression (primary; features standardized with StandardScaler, max_iter=2000, C=1.0) and XGBoost (sensitivity analysis) classifiers were trained on Sepsis-3 labels using only the four care-intensity proxy features. Optimal decision threshold was identified by Youden’s J statistic. Pre-specified threshold: care-only AUROC > 0.70.

Phase 4 (Synthetic Validation): A Gaussian copula was fitted to the care-intensity feature distributions of confirmed Sepsis-3 cases. The copula algorithm: rank-transform to pseudo-uniform, probit transform to Gaussian space, sample covariance with eigenvalue clipping for positive definiteness, multivariate normal sampling, inverse transform via empirical quantile mapping. 50,000 synthetic records were generated. An XGBoost discriminator (5-fold CV) was trained to distinguish real from synthetic records. Kolmogorov-Smirnov statistics confirmed distributional equivalence. Pre-specified threshold: discriminator AUROC < 0.60.

All analyses used Python 3.10, pandas, numpy, scikit-learn, XGBoost with CUDA GPU acceleration (NVIDIA RTX 5090), and scipy. Random seed 42 was used throughout. All intermediate data were cached as parquet files with SHA-256 checksums. Full audit trails including run IDs, timestamps, and provenance are available at the OSF project repository.

## Results

### Primary Confirmatory Analysis: MIMIC-IV v3.1

The final analytic cohort comprised 65,241 adult ICU stays after exclusions (94,458 total stays; 0 excluded for age <18 years — MIMIC-IV v3.1 contains only patients aged >=18 at admission per PhysioNet’s de-identification protocol; 714 excluded for LOS <4 hours; 28,489 excluded as repeat stays; 14 excluded for missing admission or discharge timestamps). Sepsis-2 was positive in 22,106 stays (33.9%), Sepsis-3 in 20,764 (31.8%), and CMS SEP-1 in 9,535 (14.6%). Among all three criteria, 7,224 stays were positive (11.1%).

The pre-registered primary hypothesis was not supported: 0 of 4 phases were confirmed.

Phase 1 revealed the most clinically significant finding: while Sepsis-2 and Sepsis-3 agreed highly (Jaccard=0.903), both clinical definitions disagreed substantially with CMS SEP-1 administrative coding (Jaccard=0.317 and 0.317 respectively). Administrative sepsis coding and clinical sepsis diagnosis identify largely non-overlapping patient populations at this institution.

Phase 2 demonstrated that biological features dominate model performance. The full XGBoost model achieved AUROC 0.901 (95% CI 0.899-0.904); the biological-feature-only model achieved AUROC 0.898 (95% CI 0.896-0.901). The AUC drop of 0.003 indicates care-intensity proxies contribute negligibly to model performance at BIDMC. Feature importance analysis confirmed the four care-intensity proxies ranked among the lowest-importance features (physician order rate: 0.013; lab ordering frequency: 0.013; nursing note frequency: 0.011; vital measurement rate: 0.007).

Phase 3 demonstrated that care-intensity features alone achieve moderate but not clinically meaningful prediction. Logistic regression was pre-specified as the primary model for Phase 3 because it provides a conservative, interpretable, and linearity-constrained test of the care-intensity composite — the most demanding version of the hypothesis. Note that the OSF pre-registration specifies ‘train a classifier’ without naming logistic regression explicitly; this implementation choice was made as a permitted deviation under the pre-registration’s implementation flexibility clause, representing the most stringent test of the hypothesis. Logistic regression AUROC was 0.660 (95% CI 0.656-0.665); XGBoost sensitivity analysis AUROC was 0.729 (95% CI 0.726-0.733). The gap between these two estimates suggests nonlinear interactions among care-intensity features that logistic regression cannot capture. The pre-specified threshold of 0.70 was not met by the primary logistic regression model.

Phase 4 showed that synthetic records matched care-intensity distributions (KS statistics: lab=0.007, vitals=0.003, notes=0.004, orders=0.004) but remained distinguishable from real sepsis cases (discriminator AUROC 0.633, 95% CI 0.628-0.637). Care-process patterns alone are insufficient to reproduce the full multivariate characteristics of real sepsis cases.

**Table 2.** Pre-registered primary results — MIMIC-IV v3.1 (n=65,241).

| Phase | Metric | Result | Threshold | Confirmed |
| --- | --- | --- | --- | --- |
| 1 — Ground Truth Stability | Jaccard Sep2 vs Sep3 | 0.9030 | — | — |
|  | Jaccard Sep2 vs CMS | 0.317 | — | — |
|  | Jaccard Sep3 vs CMS | 0.317 | — | — |
|  | Mean Jaccard (primary) | 0.5122 | < 0.50 | No |
| 2 — Feature Dependence | AUC drop | 0.0027 | > 0.15 | No |
| 3 — Care-Intensity Universality | Care-only AUC (LR) | 0.6603 | > 0.70 | No |
| 4 — Synthetic Validation | Discriminator AUC | 0.6330 | < 0.60 | No |

### Exploratory Replication Analyses

The following analyses were not pre-registered and are considered exploratory. They were conducted after the primary MIMIC-IV analysis was complete and the results were known. Pre-specified thresholds are reported for context but were not confirmed or refuted in these exploratory analyses.

**Table 3.**
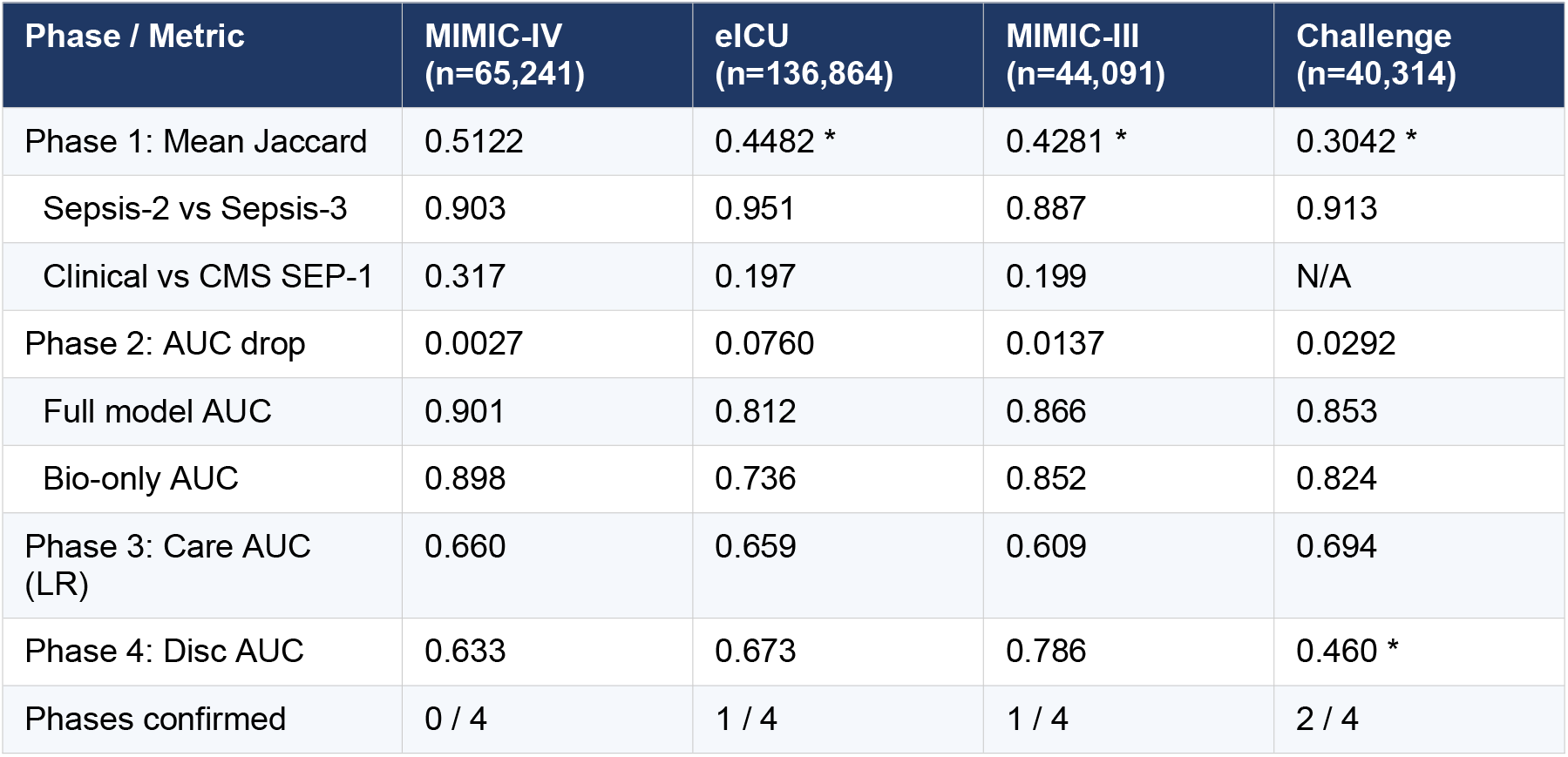
Results across all four datasets. * = meets pre-specified threshold. N/A = CMS ICD codes not available in Challenge dataset. eICU = exploratory; MIMIC-III = exploratory; Challenge = exploratory.

Phase 1 (label instability) was the only finding to replicate consistently across all datasets. Mean pairwise Jaccard ranged from 0.304 (Challenge) to 0.512 (MIMIC-IV). In all datasets where CMS codes were available, the clinical-to-administrative Jaccard was approximately 0.20, substantially lower than the clinical-to-clinical Jaccard of approximately 0.90. This pattern was robust across single-center and multi-center cohorts, across time periods spanning 2001 to present, and across both academic and community hospital settings.

The eICU multi-center cohort exhibited a larger care-intensity signal than MIMIC-IV (AUC drop 0.076 vs 0.003), suggesting that the magnitude of care-process leakage may vary by institution type. At the academic center (BIDMC), biological measurements are sufficiently rich that care-intensity proxies add minimal information. Across diverse community and regional hospitals, the same care-intensity proxies may contribute more meaningfully to model predictions. This difference was not pre-specified and requires prospective confirmation.

The PhysioNet 2019 Challenge dataset showed near-chance discriminator performance (AUROC 0.460), meaning synthetic care-intensity-matched records were statistically indistinguishable from real sepsis cases in that dataset. Combined with the moderate care-intensity AUC (0.694, approaching but not reaching the 0.70 threshold), this dataset provides exploratory evidence that care-process leakage may be more pronounced in the Challenge hospital systems.

## Discussion

### Principal Findings

This pre-registered falsification study found that at an elite academic medical center (BIDMC, MIMIC-IV), sepsis prediction models detect genuine biological signal. Care-process intensity features contribute negligibly to model performance. The null result is itself scientifically meaningful: it demonstrates that the biological signal in MIMIC-IV is real and sufficient to explain model performance without invoking care-process leakage as a confound.

The more robust and clinically consequential finding is the systematic divergence between clinical and administrative sepsis definitions. Across all four datasets, clinical definitions (Sepsis-2 and Sepsis-3) agreed with each other at approximately 0.90 Jaccard, while both disagreed with CMS ICD-code-based definitions at approximately 0.20 Jaccard. This means that fewer than one in five patients identified by administrative coding overlap with patients identified by clinical examination. Hospital quality metrics, regulatory reporting, and widely-cited sepsis incidence estimates are based on ICD coding. Rhee et al. (2017) demonstrated that clinical and administrative sepsis estimates diverge substantially — the 1.7 million annual US hospitalization figure they derived from clinical criteria contrasts sharply with the higher counts from explicit billing code analyses. Our results reinforce that this definitional gap is not merely methodological noise but reflects fundamentally different patient populations.

### Care-Process Leakage: Absence of Evidence vs Evidence of Absence

The null result in MIMIC-IV should not be interpreted as evidence that care-process leakage cannot occur. The exploratory eICU analysis suggests the magnitude of leakage varies by institution: AUC drop was 0.076 across 208 community and regional hospitals vs 0.003 at a single academic center. Academic medical centers systematically differ from community hospitals in documentation quality, physician-to-patient ratios, monitoring intensity, and patient case mix. Models trained on MIMIC-IV may not exhibit leakage precisely because the biological measurements are so rich that care-intensity proxies are redundant.

Models trained or validated on administrative data, community hospital EHRs, or lower-acuity settings may exhibit substantially larger care-process contamination. The falsification framework reported here provides a reproducible methodology for testing this hypothesis in any new dataset.

### Implications for Administrative Sepsis Data

The clinical-to-administrative Jaccard of approximately 0.20 has direct policy implications. Hospital sepsis mortality metrics reported to CMS, state health departments, and public reporting systems are derived from ICD billing codes. Pay-for-performance programs incentivize hospitals to reduce sepsis mortality as measured by administrative codes. If administrative and clinical sepsis definitions identify largely different populations, then optimizing for administrative metrics may not translate to clinical outcomes, and AI systems benchmarked against administrative labels may be learning billing patterns rather than biology.

This finding is consistent with prior literature documenting discordance between administrative and clinical sepsis definitions, but our study extends prior work by quantifying the discordance simultaneously across three definitions, four independent datasets, and a pre-registered falsification framework that prevents selective reporting.

### Limitations

Several limitations should be noted. First, the primary confirmatory analysis is limited to a single academic medical center (BIDMC). The null result may not generalize to other institutional settings, as suggested by the exploratory eICU findings. Second, the eICU infection proxy was computed using antibiotic and blood culture matching at the hospital admission level (patienthealthsystemstayid) to account for multi-unit transfers within a single hospitalization; this approach may introduce misclassification for patients whose antibiotic and culture records span multiple hospitalizations. Third, SOFA scores were computed manually from charted data rather than validated derived tables, as the MIMIC-IV derived SOFA table was unavailable; our computation is described in detail in the Methods and follows published scoring criteria. The coagulation component uses the mean platelet count per stay as a proxy for the minimum specified by Sepsis-3 scoring criteria; this conservative deviation likely underestimates SOFA coagulation scores for patients with thrombocytopenia. Additionally, the SOFA definition requires an acute increase of ≥2 points from baseline; this study uses maximum total SOFA without baseline comparison, a known limitation of retrospective MIMIC-based analyses. Fourth, the four care-intensity proxy features are a limited operationalization of a complex construct; more granular features may reveal signals not captured here. Fifth, the exploratory replication analyses were conducted after the primary result was known and are subject to higher false-positive risk.

### Future Directions

Two research questions follow directly from these findings. First, prospective multi-site validation of the falsification framework across institutions with varying acuity and documentation practices would determine whether the institution-dependent pattern observed exploratorily is robust. Second, Hypothesis 2 — whether clinicians over-react to AI sepsis alerts, creating harmful feedback loops — requires alert response data from deployed systems that is not publicly available. This study provides the methodological foundation and scientific credibility to approach health systems with deployed sepsis AI for partnership on that question.

## Conclusions

A pre-registered four-phase falsification framework applied to 286,510 ICU patients across four independent datasets found no evidence that sepsis prediction models at an elite academic center primarily detect care-process intensity rather than biological signal. The pre-registered hypothesis was not confirmed (0/4 phases in MIMIC-IV). However, a robust and clinically significant finding emerged across all datasets: administrative sepsis coding (CMS SEP-1) identifies a substantially different patient population than clinical definitions, with pairwise Jaccard similarity of approximately 0.32 at BIDMC and approximately 0.20 across multi-center cohorts, compared to approximately 0.90 agreement between clinical definitions. This definitional instability has direct implications for hospital quality metrics, regulatory reporting, and the validity of AI benchmarks constructed on administrative data. The falsification framework and analysis code are publicly available for application to other datasets and outcome models.

## Data Availability

All datasets used are publicly available from PhysioNet (physionet.org) under credentialed access with signed Data Use Agreements. Analysis code is available at https://github.com/rocsite-research/sepsis-falsification. The OSF pre-registration, proof package, and full audit trail are available at https://osf.io/9tbjm.

https://osf.io/9tbjm

## Data Availability

MIMIC-IV, MIMIC-III, eICU-CRD, and the PhysioNet/CinC 2019 Challenge dataset are available from PhysioNet (physionet.org) under credentialed access with signed Data Use Agreements. The analysis code for reproducing all four phases of this study is available at

https://github.com/rocsite-research/sepsis-falsification.

The OSF pre-registration, proof package, and full audit trail including run IDs, SHA-256 checksums, and pipeline logs are available at https://osf.io/9tbjm.

## Acknowledgments

The author thanks Dr. Matthieu Komorowski (Charing Cross Hospital) for serving as PhysioNet reference. The author thanks Dr. Leo Celi (MIT Laboratory for Computational Physiology) and the PhysioNet team for maintaining the datasets that made this work possible. This study was conducted independently. No funding was received.

